# A Sustainable Development Goal framework to guide multisectoral action on NAFLD through a societal approach

**DOI:** 10.1101/2021.08.02.21261464

**Authors:** Jeffrey V Lazarus, Henry E Mark, Massimo Colombo, Sandro Demaio, John F Dillon, Jacob George, Hannes Hagström, Samantha Hocking, Nancy Lee, Mark J Nieuwenhuijsen, Mary E Rinella, Manuel Romero-Gomez, Joan B Soriano, Jörn M Schattenberg, Frank Tacke, Emmanuel A. Tsochatzis, Luca Valenti, Shira Zelber-Sagi, M. Ashworth Dirac, Terry T-K Huang

## Abstract

Non-alcoholic fatty liver disease (NAFLD) is a highly prevalent condition that requires a comprehensive and coordinated response across sectors and disciplines. In the absence of a multisectoral framework, we developed a NAFLD-Sustainable Development Goals (SDG) framework to converge thinking about the design and delivery of NAFLD public health responses. A multidisciplinary group identified SDG targets and indicators for inclusion in the NAFLD-SDG framework through a two-stage process. Firstly, a core team of three researchers independently reviewed the 169 SDG targets and 231 unique indicators proposed by the Inter-Agency and Expert Group on SDG to select a shortlist. Over two Delphi rounds, a multidisciplinary group of 12 experts selected which of the shortlisted targets and indicators to include in the NAFLD-SDG framework. Respondents also provided written feedback on their selection. Targets and indicators with 75% or greater agreement were included in the final NAFLD-SDG framework. The final framework comprises 16 targets–representing 9% of all SDG targets and 62% (16/26) of the shortlisted targets–and seven indicators, accounting for 50% (7/14) of the shortlisted indicators and 3% of all SDG indicators. The selected targets and indicators cover a broad range of factors, from health, food and nutrition to education, the economy and the built environment. Addressing the challenge of NAFLD will require re-envisioning the liver health landscape, with a greater focus on joined-up systems thinking and action. The NAFLD-SDG framework can help guide this process, including by outlining the key stakeholders with whom the liver health community needs to engage.

## Introduction

There is increasing recognition of the need to view complex health issues as systems problems, requiring multidisciplinary and multisectoral responses.^1^ Non-alcoholic fatty liver disease (NAFLD) is a potentially serious liver condition that affects close to 1 in 4 adults globally,^2,3^ causing substantial impairment to health-related quality of life^4^ and significant healthcare costs and economic loss.^5^ Despite the scale of the challenge, NAFLD remains largely unknown outside the field of hepatology and public health response have been weak and fragmented.^6^

Part of a multisystem disease, NAFLD shares a complex bidirectional relationship with components of the metabolic syndrome. NAFLD is a leading cause of cirrhosis^7^ and contributes to cardiovascular disease, type 2 diabetes and non-hepatic cancer morbidity and mortality.^8,9^ NAFLD is strongly–but not exclusively–associated with obesity^10^ and is still common in non-obese individuals.^11^

NAFLD shares common risk factors with other highly prevalent non-communicable diseases (NCDs), namely those related to food systems and the lived environment. As with NCDs more broadly, addressing NAFLD will require a comprehensive, cohesive, and coordinated response that synergistically acts upon the immediate, underlying and basic influences of disease. The lack of joined-up efforts, including a comprehensive multisectoral framework for addressing NAFLD, is a barrier to the design and delivery of public health responses to this disease.

The Sustainable Development Goals (SDGs) serve as the mainstay of the 2030 Agenda for Sustainable Development with clear priorities, from reducing social and economic inequalities to improving nutrition, health and education.^12^ The 17 goals and 169 targets that make up the SDGs provide a blueprint for cross-sectoral collaboration and action. Previous studies have used the SDGs to develop a conceptual framework for informing policy approaches on sustainable development and urban health^13^ and to highlight the importance of addressing obesity for achieving the sustainable development agenda.^14^

We set out to develop a NAFLD-SDG framework to help conceptualise the design and delivery of a comprehensive, multisectoral public health response to NAFLD.

## Methods

The NAFLD conceptual model developed by Lazarus et al^*15*^ outlines the basic, underlying and direct influences that contribute to the development of the disease. We grouped elements of this model into six domains: 1. economic, political and social context; 2. health; 3. nutrition and food environment; 4. welfare and social services; 5. education; and 6. lived environment, in order to link the elements of the model to specific SDG targets and indicators (Supplementary Table 1).

The SDGs consist of 17 goals, followed by targets within each goal and indicators within each target. Targets are specific objectives while indicators serve as quantifiable metrics. The selection of targets and indicators for inclusion in the NAFLD-SDG framework followed a two-stage process. Firstly, a core team of three researchers (JVL, HEM and TTKH) independently reviewed the 169 SDG targets and 231 unique indicators proposed by the Inter-Agency and Expert Group on SDG,^16^ selecting targets and indicators directly or indirectly linked to one of the six domains. The researchers unanimously agreed upon the shortlist. Not all indicators within the selected targets were included as some accompanying indicators were insufficiently specific or relevant to NAFLD. Conversely, if an indicator was included, the target under which it fell had to be included.

The second stage of the selection followed a Delphi process over two rounds of voting. In the first round, a multidisciplinary group of 12 experts were invited to select which of the shortlisted targets and indicators to include in the NAFLD-SDG framework. Respondents also provided written feedback on their selection. Nine of the 12 experts (75%) completed the round 1 survey. For round 2, the survey included a summary of the written feedback provided by respondents in round 1. Targets which received less than 25% agreement in round 1 were removed (n=1). In the second round of voting, all 15 individuals completed the survey (the 12 experts and three core team members). The core group members who led the analysis completed the round 2 survey prior to other respondents so as not to be influenced by the selection of others. Targets and indicators with 75% or greater agreement were included in the final NAFLD-SDG framework.

## Results

Table 1 reports the final level of consensus achieved for each of the shortlisted targets and indicators. A total of 16 targets across eight SDGs were included in the framework, representing 9% of all SDG targets and 62% (16/26) of the shortlisted targets. Seven indicators across five SDGs were also included in the framework, accounting for 50% (7/14) of the shortlisted indicators and 3% of all SDG indicators. (Table 3, Figure 2).

**Table 1:**
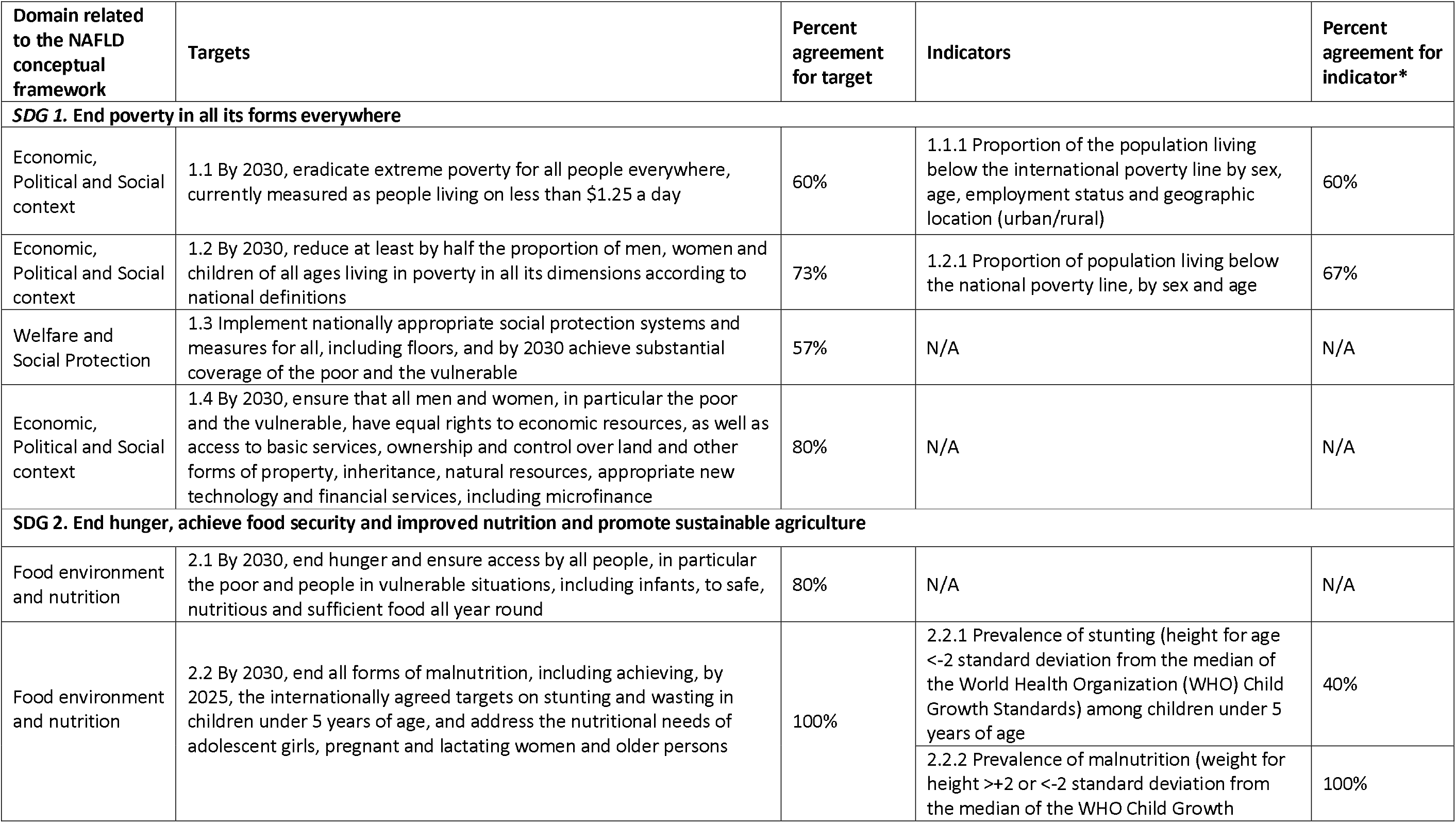
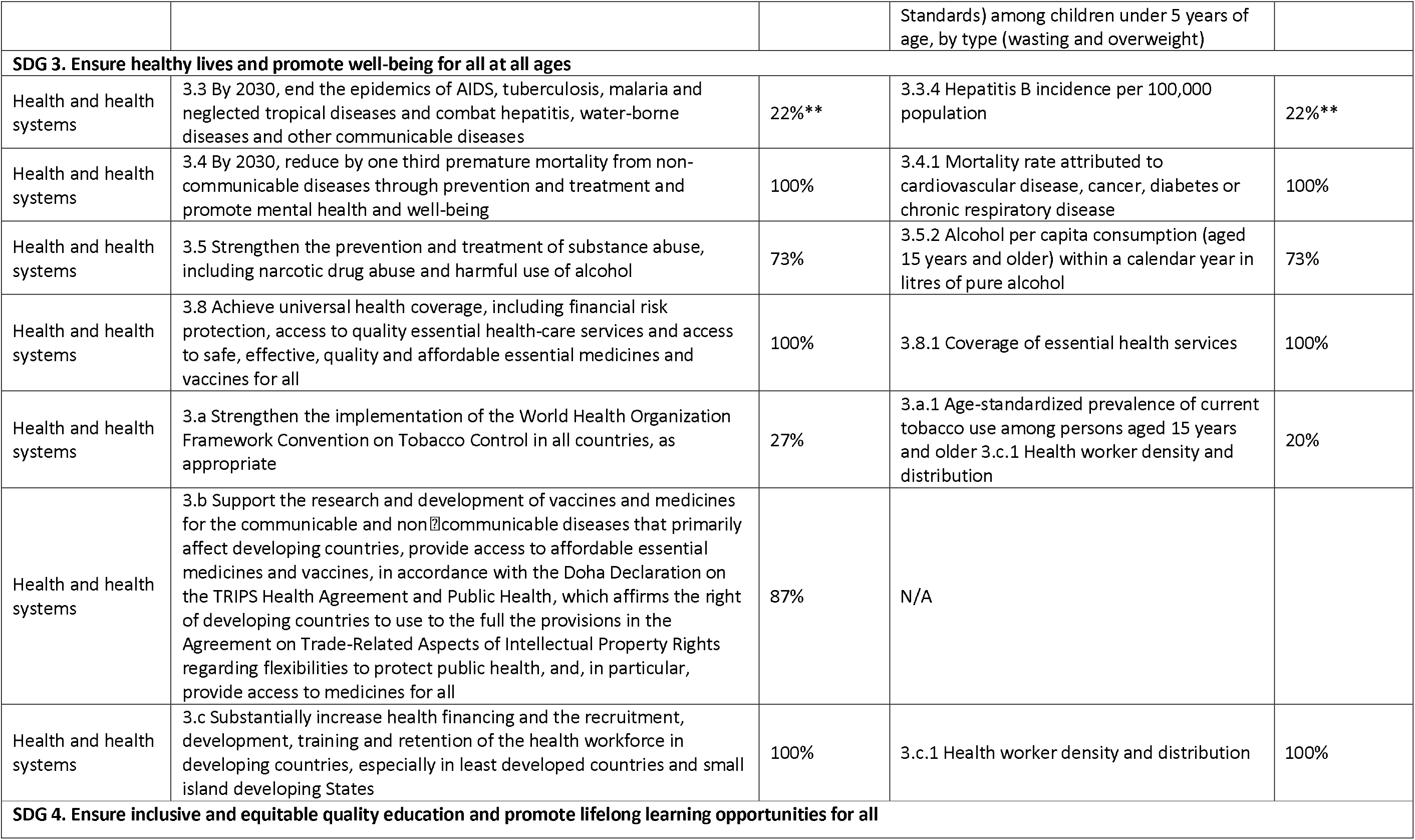
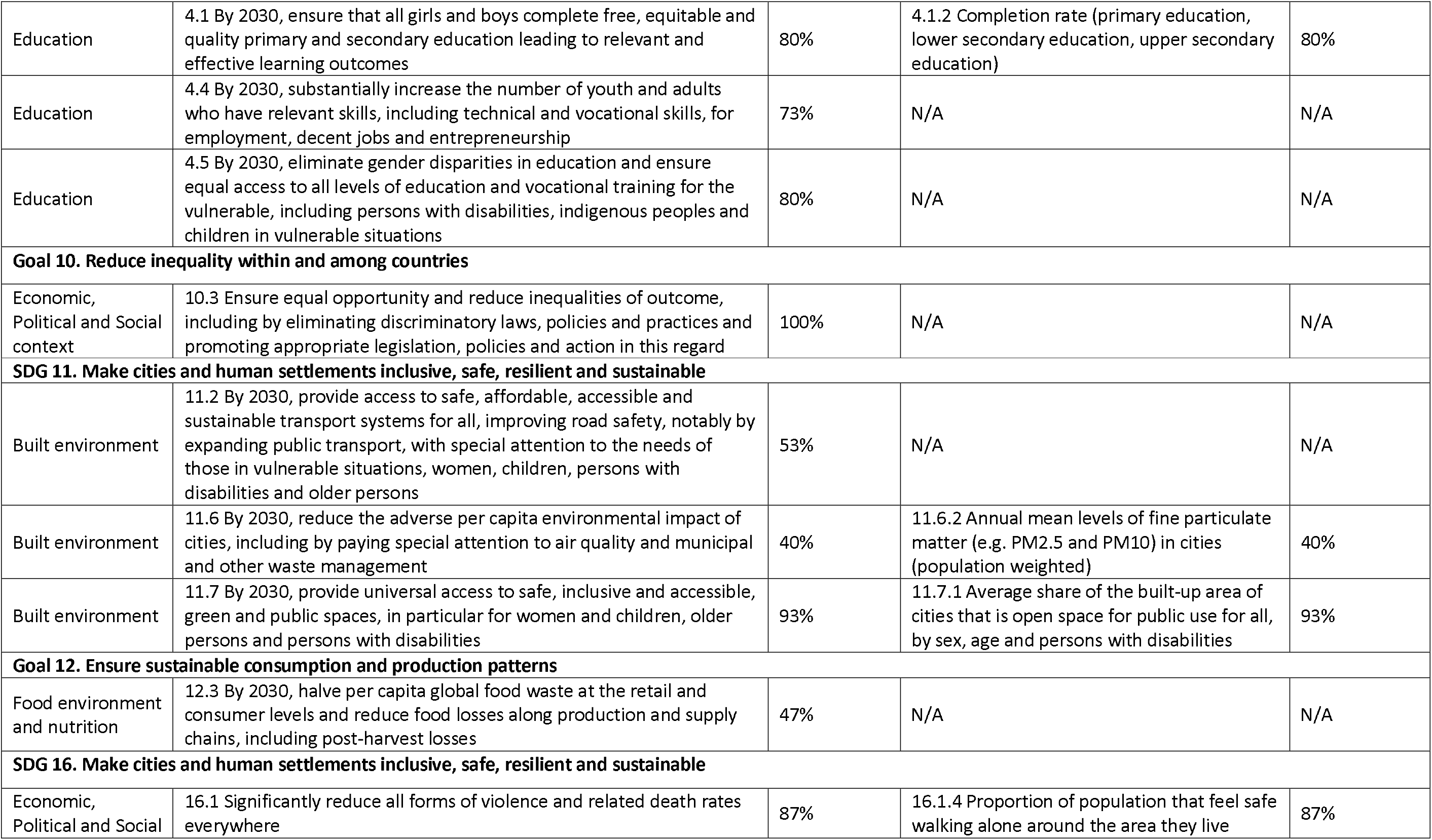
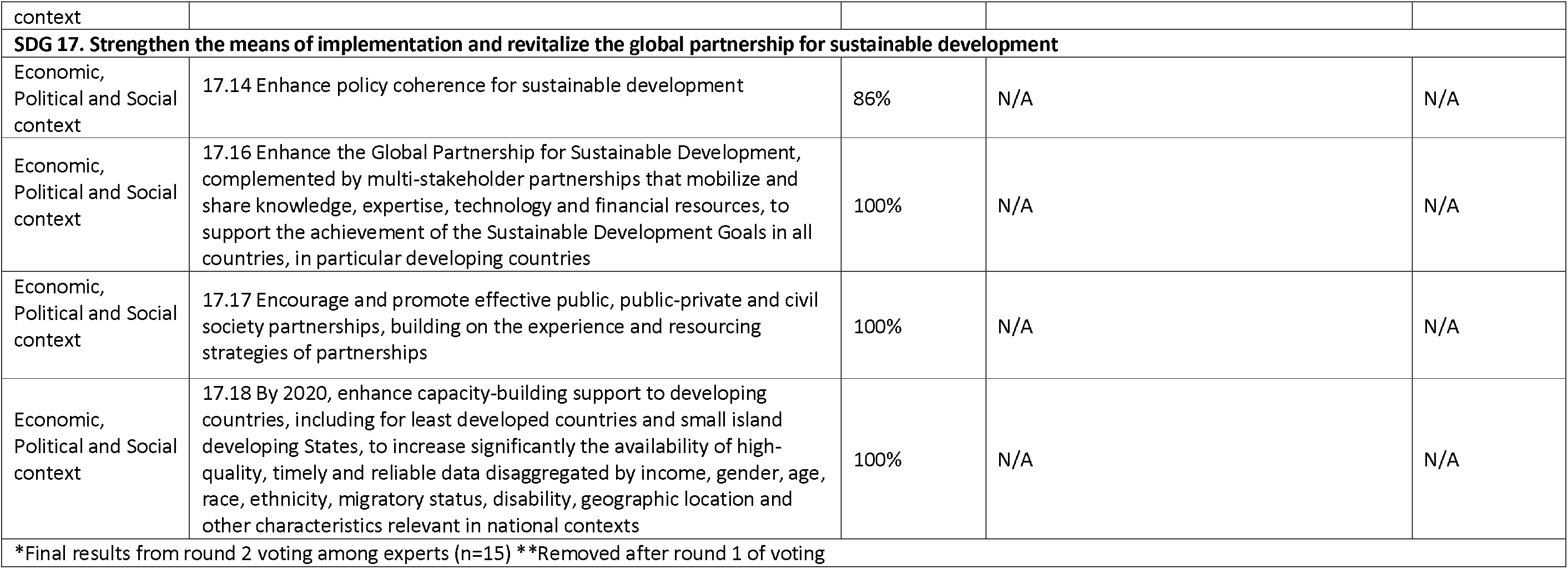
Sustainable Development Goals and associated targets and indicators shortlisted for inclusion in the SDG-NAFLD framework and final level of consensus

| Domain related to the NAFLD conceptual framework | Targets | Percent agreement for target | Indicators | Percent agreement for indicator* |
| --- | --- | --- | --- | --- |
| **SDG 1. End poverty in all its forms everywhere** |  |  |  |  |
| Economic, Political and Social context | 1.1 By 2030, eradicate extreme poverty for all people everywhere, currently measured as people living on less than \$1.25 a day | 60% | 1.1.1 Proportion of the population living below the international poverty line by sex, age, employment status and geographic location (urban/rural) | 60% |
| Economic, Political and Social context | 1.2 By 2030, reduce at least by half the proportion of men, women and children of all ages living in poverty in all its dimensions according to national definitions | 73% | 1.2.1 Proportion of population living below the national poverty line, by sex and age | 67% |
| Welfare and Social Protection | 1.3 Implement nationally appropriate social protection systems and measures for all, including floors, and by 2030 achieve substantial coverage of the poor and the vulnerable | 57% | N/A | N/A |
| Economic, Political and Social context | 1.4 By 2030, ensure that all men and women, in particular the poor and the vulnerable, have equal rights to economic resources, as well as access to basic services, ownership and control over land and other forms of property, inheritance, natural resources, appropriate new technology and financial services, including microfinance | 80% | N/A | N/A |
| **SDG 2. End hunger, achieve food security and improved nutrition and promote sustainable agriculture** |  |  |  |  |
| Food environment and nutrition | 2.1 By 2030, end hunger and ensure access by all people, in particular the poor and people in vulnerable situations, including infants, to safe, nutritious and sufficient food all year round | 80% | N/A | N/A |
| Food environment and nutrition | 2.2 By 2030, end all forms of malnutrition, including achieving, by 2025, the internationally agreed targets on stunting and wasting in children under 5 years of age, and address the nutritional needs of adolescent girls, pregnant and lactating women and older persons | 100% | 2.2.1 Prevalence of stunting (height for age <-2 standard deviation from the median of the World Health Organization (WHO) Child Growth Standards) among children under 5 years of age | 40% |
|  |  |  | 2.2.2 Prevalence of malnutrition (weight for height >+2 or <-2 standard deviation from the median of the WHO Child Growth | 100% |
|  |  |  | Standards) among children under 5 years of age, by type (wasting and overweight) |  |
| **SDG 3. Ensure healthy lives and promote well-being for all at all ages** |  |  |  |  |
| Health and health systems | 3.3 By 2030, end the epidemics of AIDS, tuberculosis, malaria and neglected tropical diseases and combat hepatitis, water-borne diseases and other communicable diseases | 22%** | 3.3.4 Hepatitis B incidence per 100,000 population | 22%** |
| Health and health systems | 3.4 By 2030, reduce by one third premature mortality from non-communicable diseases through prevention and treatment and promote mental health and well-being | 100% | 3.4.1 Mortality rate attributed to cardiovascular disease, cancer, diabetes or chronic respiratory disease | 100% |
| Health and health systems | 3.5 Strengthen the prevention and treatment of substance abuse, including narcotic drug abuse and harmful use of alcohol | 73% | 3.5.2 Alcohol per capita consumption (aged 15 years and older) within a calendar year in litres of pure alcohol | 73% |
| Health and health systems | 3.8 Achieve universal health coverage, including financial risk protection, access to quality essential health-care services and access to safe, effective, quality and affordable essential medicines and vaccines for all | 100% | 3.8.1 Coverage of essential health services | 100% |
| Health and health systems | 3.a Strengthen the implementation of the World Health Organization Framework Convention on Tobacco Control in all countries, as appropriate | 27% | 3.a.1 Age-standardized prevalence of current tobacco use among persons aged 15 years and older<br>3.c.1 Health worker density and distribution | 20% |
| Health and health systems | 3.b Support the research and development of vaccines and medicines for the communicable and non-communicable diseases that primarily affect developing countries, provide access to affordable essential medicines and vaccines, in accordance with the Doha Declaration on the TRIPS Health Agreement and Public Health, which affirms the right of developing countries to use to the full the provisions in the Agreement on Trade-Related Aspects of Intellectual Property Rights regarding flexibilities to protect public health, and, in particular, provide access to medicines for all | 87% | N/A |  |
| Health and health systems | 3.c Substantially increase health financing and the recruitment, development, training and retention of the health workforce in developing countries, especially in least developed countries and small island developing States | 100% | 3.c.1 Health worker density and distribution | 100% |
| **SDG 4. Ensure inclusive and equitable quality education and promote lifelong learning opportunities for all** |  |  |  |  |
| Education | 4.1 By 2030, ensure that all girls and boys complete free, equitable and quality primary and secondary education leading to relevant and effective learning outcomes | 80% | 4.1.2 Completion rate (primary education, lower secondary education, upper secondary education) | 80% |
| Education | 4.4 By 2030, substantially increase the number of youth and adults who have relevant skills, including technical and vocational skills, for employment, decent jobs and entrepreneurship | 73% | N/A | N/A |
| Education | 4.5 By 2030, eliminate gender disparities in education and ensure equal access to all levels of education and vocational training for the vulnerable, including persons with disabilities, indigenous peoples and children in vulnerable situations | 80% | N/A | N/A |
| **Goal 10. Reduce inequality within and among countries** |  |  |  |  |
| Economic, Political and Social context | 10.3 Ensure equal opportunity and reduce inequalities of outcome, including by eliminating discriminatory laws, policies and practices and promoting appropriate legislation, policies and action in this regard | 100% | N/A | N/A |
| **SDG 11. Make cities and human settlements inclusive, safe, resilient and sustainable** |  |  |  |  |
| Built environment | 11.2 By 2030, provide access to safe, affordable, accessible and sustainable transport systems for all, improving road safety, notably by expanding public transport, with special attention to the needs of those in vulnerable situations, women, children, persons with disabilities and older persons | 53% | N/A | N/A |
| Built environment | 11.6 By 2030, reduce the adverse per capita environmental impact of cities, including by paying special attention to air quality and municipal and other waste management | 40% | 11.6.2 Annual mean levels of fine particulate matter (e.g. PM2.5 and PM10) in cities (population weighted) | 40% |
| Built environment | 11.7 By 2030, provide universal access to safe, inclusive and accessible, green and public spaces, in particular for women and children, older persons and persons with disabilities | 93% | 11.7.1 Average share of the built-up area of cities that is open space for public use for all, by sex, age and persons with disabilities | 93% |
| **Goal 12. Ensure sustainable consumption and production patterns** |  |  |  |  |
| Food environment and nutrition | 12.3 By 2030, halve per capita global food waste at the retail and consumer levels and reduce food losses along production and supply chains, including post-harvest losses | 47% | N/A | N/A |
| **SDG 16. Make cities and human settlements inclusive, safe, resilient and sustainable** |  |  |  |  |
| Economic, Political and Social | 16.1 Significantly reduce all forms of violence and related death rates everywhere | 87% | 16.1.4 Proportion of population that feel safe walking alone around the area they live | 87% |
| context |  |  |  |  |
| **SDG 17. Strengthen the means of implementation and revitalize the global partnership for sustainable development** |  |  |  |  |
| Economic, Political and Social context | 17.14 Enhance policy coherence for sustainable development | 86% | N/A | N/A |
| Economic, Political and Social context | 17.16 Enhance the Global Partnership for Sustainable Development, complemented by multi-stakeholder partnerships that mobilize and share knowledge, expertise, technology and financial resources, to support the achievement of the Sustainable Development Goals in all countries, in particular developing countries | 100% | N/A | N/A |
| Economic, Political and Social context | 17.17 Encourage and promote effective public, public-private and civil society partnerships, building on the experience and resourcing strategies of partnerships | 100% | N/A | N/A |
| Economic, Political and Social context | 17.18 By 2020, enhance capacity-building support to developing countries, including for least developed countries and small island developing States, to increase significantly the availability of high-quality, timely and reliable data disaggregated by income, gender, age, race, ethnicity, migratory status, disability, geographic location and other characteristics relevant in national contexts | 100% | N/A | N/A |
| *Final results from round 2 voting among experts (n=15) **Removed after round 1 of voting |  |  |  |  |

Below, we summarise the targets and indicators included within each of the six domains. Figure 2 provides a graphical representation of the NAFLD-SDG framework and differentiates the targets into those having a direct or indirect influence on NAFLD. The indirect targets are generally upstream factors that have the potential to influence NAFLD via their impact on downstream elements.

### Economic, political and social context

Seven targets across four SDGs were included in the economic, political and social context domain. These seven targets represent the distal factors that have a downstream influence on the development of NAFLD.

SDG 1.4, related to equal access to economic resources and basic services, was the only SDG 1 target included in the framework. Shortlisted targets and indicators related to poverty eradication and social protection systems did not meet the threshold for inclusion. While poverty eradication is of central importance to the global development agenda, this has a relatively indirect influence on the development of NAFLD. Economic growth has a complex relationship with metabolic diseases, including NAFLD. This is in part due to the proliferation of unhealthy diets and sedentary behaviour that are associated with increasingly westernised lifestyles. In low-income settings, increasing socio-economic status may be accompanied by changes in diet and lifestyle associated with the risk of developing NAFLD. Through a NAFLD lens, focus should be placed on reducing inequalities, and improving health literacy while ensuring that economic development is accompanied by appropriate public health measures and improved access to health services. SDG 10.3, which is included in the framework, specifically relates to reducing inequities of outcomes, including health.

SDG 16.1 on reducing violence was included in the framework along with indicator 16.1.4 on public perception about feeling safe when walking near home. While the association to NAFLD is indirect, this target and its related indicator provide important insights on people’s ability to freely and safely exercise near their homes, which is a prerequisite for increasing levels of physical activity.

Four SDG 17 targets addressing cross-cutting factors around governance, partnerships and knowledge were included in the framework. While indirectly related to NAFLD, these factors provide a foundation for integrated and collaborative approaches to sustainable development, which are central to addressing major global health challenges.

### Food environment and nutrition

Target 2.1 (ending hunger), target 2.2 (ending all forms of malnutrition), and indicator 2.2.2 (prevalence of malnutrition) were included in the framework. Ending hunger and childhood undernutrition can have a downstream preventive impact on NAFLD and other metabolic conditions given how early life undernutrition is associated with chronic disease in later life. Overweight and obesity are major risk factors for NAFLD and tackling obesity in all ages will be critical for efforts to prevent and manage the disease. The liver health community should directly engage with actors in the obesity space to identify areas for collaboration from health system responses to broader public health measures.

### Health and health systems

Four of the SDG3 targets and three indicators were included in the framework. Three of these targets provide mechanisms for addressing NAFLD, from increasing access to healthcare services through universal health coverage (3.8) and growing and training the health workforce (3c), to research and development of medicines for NCDs (3b). Within these targets, the liver health community can lead on a range of specific activities, from advancing the understanding of how best to deliver health services to people living with chronic metabolic conditions, to increasing the awareness, knowledge and capacity of the health workforce to meet the needs of people living with NAFLD.

The remaining target (3.4) is focused on reducing premature mortality from NCDs by one-third. NAFLD contributes directly to this target as non-alcoholic steatohepatitis (NASH)–the more serious form of NAFLD–is an increasing cause of end-stage liver disease. NAFLD also has an indirect impact, being an early sign of the metabolic dysregulation that leads to cardiovascular disease – a leading cause of death in people living with NAFLD. Addressing NAFLD will be important for the attainment of target 3.4, providing further rationale for addressing this condition as part of broader efforts to prevent and manage NCDs.

### Education

Two education targets (4.1 primary and secondary schooling completion and 4.5 reducing gender disparities in education) and one indicator (4.1.2 primary and secondary school completion rates) were included in the framework. Equitable and consistent access to education is indirectly related to NAFLD, with childhood education level being associated with health outcomes in adulthood, including NCDs. While the liver health community has no direct involvement in schooling, we can support wider calls within the global public health community to ensure equal access and attainment of basic schooling for all, recognising this as a foundation for health and well-being.

### Built environment

The built environment can have a marked effect on health through its role in recreation, exercise and mental well-being. SDG 11.7 (universal access to safe, inclusive and accessible, green and public spaces) and its associated indicator 11.7.1 (share of open space for public use in cities) were included in the framework. Access to green spaces was selected as a proxy for the liveability of urban spaces. Related to green spaces are the issues of air pollution and re-pedestrianizing streets and improving the walkability and cyclability of roads also fall within this domain.

## Discussion

Despite affecting 23-25% of the global adult population,^2,3^ NAFLD remains largely unknown within the global public health community, with little appreciation of the relationship between NAFLD and other highly prevalent diseases and the need for multisectoral responses to address the challenge. We applied a NAFLD lens to the SDGs, creating a framework to help stakeholders envisage the comprehensive, multisectoral strategies required to address NAFLD.

The NAFLD conceptual model that provided the initial starting point for this paper outlined basic influences at the socio-economic and political levels, underlying influences related to social systems and direct influences such as nutrition. Critically, because the SDGs encompass all of these levels of influence and are used worldwide to guide national and international action, they provide a foundational framework for considering the interdependence of different sectors and systems. The NAFLD-SDG framework can serve a number of purposes for the liver health and public health communities. Firstly, at a conceptual level, it can support a shift in thinking around NAFLD, from a narrow understanding of the immediate causes of the disease to considering the broader systems and structures that have enabled this to become the most prevalent liver disease in history. Secondly, it can help stakeholders map out strategic priorities across different sectors, including where the liver health community needs to lead the agenda, and the many more areas where the community needs to engage in and support ongoing efforts.

Our framework outlines how NAFLD is implicitly captured in the SDGs through its association with many existing goals, targets and indicators. Nonetheless, there is currently no explicit NAFLD target or indicator; this presents several challenges. The SDGs are used to inform decisions at national and global levels, including which areas to prioritise and fund. NAFLD remains largely obscure outside the liver heath community; specific mention of the disease within the SDGs would be a powerful way to increase awareness and drive action. We would propose that NAFLD be explicitly mentioned in indicator 3.4.1 on NCD mortality rates; this indicator currently includes cardiovascular disease, cancer, diabetes and chronic respiratory disease.

### Re-visioning the liver health landscape

Historically, the liver health community has rarely considered multidisciplinary approaches, focusing instead on increasing access to treatment. However, this began to change with the advent of new curative treatments for viral hepatitis C in 2013. Despite the availability of new treatments, the prevalence of hepatitis C infection did not decline.^17^ It became clear that a whole-of-systems approach would be needed to address hepatitis C, including engaging with at-risk populations, patient groups, addiction specialists and general practitioners, both to prevent infection and to diagnose and treat it.^18^

The liver health field must now further expand its horizons to look beyond the health sector as it seeks to address the challenge of NAFLD. This will require a re-visioning of the liver health landscape for the years to come and embracing systems thinking. The liver health community can draw inspiration from other fields that have had some success in recent years, including the obesity field. In the past two decades, popular thinking on obesity has shifted from a focus on individual-level factors underlying energy imbalance to considering biological, social, environmental and policy drivers of health behaviours and outcomes.^19^ More recently, research has gone one step further to emphasize the interconnections across these levels of influence in an integrated, systems approach.^20^ Such an approach calls for coordinated actions from all stakeholders and requires improving policies and practices across multiple sectors as well as shifting social norms on health.^21^ For NAFLD, we need a similar paradigm shift. The NAFLD-SDG framework provides the guidance for this.

### Integrating NAFLD within the NCD agenda and ongoing public health efforts

At both a public health and clinical management level, there is substantial overlap in the measures required to address NAFLD and the other major NCDs. However, NAFLD is not mentioned by name in almost any key global or national NCD strategy, most notably the WHO Global Action Plan on the Prevention and Control of NCDs.^22^ At a health system level, chronic disease management is driving the need for the reorientation of health systems away from siloed disease-centric models to multidisciplinary patient-centred care. The liver community, through collaboration with others working on metabolic disease management, can help lead this process in the years to come.

Given the relationship between NAFLD, obesity and other highly prevalent NCDs, namely type 2 diabetes and heart disease, integration of NAFLD within the NCD agenda makes sense from both strategic and operational perspectives. By doing so, the liver heath community can become a powerful ally for the NCD community. Significant efforts are underway to establish alliances across organizations dedicated to NCDs, both to increase political clout and drive multisectoral action. It is now time for the liver community to join in these efforts.

### Strengths and limitations

Our paper has several strengths and limitations. The selection of targets and indicators is a somewhat subjective process. In the shortlisting phase, we reduced the risk of individual bias by having three researchers independently review all these before unanimously agreeing the targets and indicators to be put forward for voting. The diversity of the panellists also has a substantial impact on the outcome of the Delphi process. To ensure that a breadth of perspectives was considered, we engaged experts from a range of disciplines, from liver health and NCDs, to food and nutrition, and the SDGs. One of the major strengths of the paper lies in the use of the SDGs. Developed through a collaborative multi-stakeholder process, the SDGs are a widely known and utilised framework, thus making our findings applicable to a wide audience.

## Conclusion

The NAFLD-SDG framework can act as a strategic advocacy tool to build the case for closer collaboration within and between sectors seeking to address NAFLD and other NCDs. The liver health community needs to champion the cause of NAFLD and should seek to engage with existing efforts within other relevant sectors. A NAFLD public health roadmap, developed through broad engagement with key stakeholders, can help guide this process and translate the NAFLD-SDG framework from concept to action.

**Figure 1.**
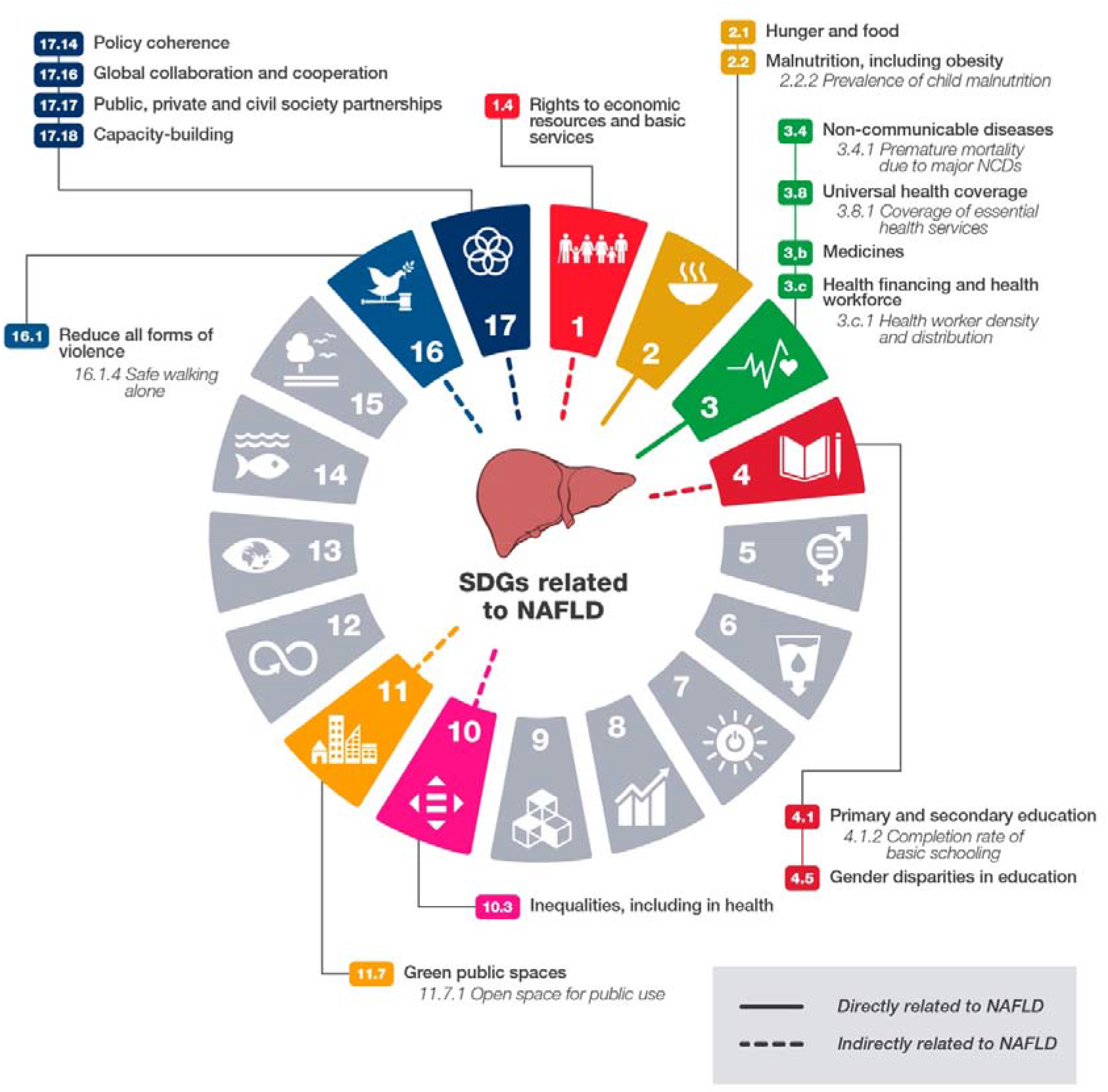
NAFLD-SDG framework.

## Data Availability

Data are available from the corresponding author upon request.

## Author Contributions

JVL and HEM conceptualised the study. JVL, HEM and TTKH shortlisted the targets and indicators. JVL, HEM, SD, JG, SH, NL, MER, MRG, JBS, JMS, LV, SZS, MAD and TTKH participated in the Delphi rounds. HEM and JVL led the data collection and analysis. HEM, JVL and TTKH developed the first draft of the manuscript, all authors provided input and feedback on the manuscript drafts and reviewed the final version prior to submission.

## Acknowledgements

This work was supported by the EASL International Liver Foundation who acknowledges funding from Intercept Pharmaceutics, as well as Bristol Myers Squibb and Merck Sharp & Dohme. The funders had no role in the study design; in the collection, analysis, and interpretation of data; in the writing of the report; and in the decision to submit the paper for publication.

