## Supplementary Table 1 for "A Sustainable Development Goal framework to guide multisectoral action on NAFLD through a societal approach"

**Supplementary Table 1:** Domains related to the NAFLD conceptual framework

| **Domain** | **Examples** | **Description** |
| --- | --- | --- |
| ***Economics, Political and Social context*** | Economic inequality | Level of poverty and economic inequality |
|  | Political and social inequalities | Inclusion of all groups in economic and political decision-making |
| **Health** | Health systems | Access to quality, affordable health services for NAFLD patients and those suffering from common-comorbid conditions |
|  | Health promotion | Prevention of NAFLD and common co-morbidities (e.g., through promotion of healthy lifestyles), promotion of mental wellbeing and positive health-seeking behaviours |
| **Food** | Food environment | Availability of and access to a safe and nutritious diet |
|  | Malnutrition | Prevention of all forms of malnutrition, including overweight and obesity |
|  | Marketing and promotion | Promotional activities related to the marketing of unhealthy foods |
| **Welfare and social protection** | Social protection | Access to adequate social protection for vulnerable population groups |
| **Education systems** | Basic education | Access to quality, affordable basic education regardless of social or economic context (e.g. gender, socio-economic status) |
|  | Learning outcomes | Promotion of high levels of literacy and numeracy |
| **Lived environment** | Access to green spaces | Access to green spaces and recreational areas for exercise |
|  | Basic public services | Availability of and access to basic public services including safe water and sanitation |
|  | Pollution | Levels of air pollution |
